# Negative impact of the COVID-19 pandemic on sleep quantitative parameters, quality, and circadian alignment: Implications for psychological well-being and emotional regulation

**DOI:** 10.1101/2020.07.09.20149138

**Authors:** Mohammad Ali Salehinejad, Maryam Majidinezhad, Elham Ghanavati, Sahar Kouestanian, Carmelo M. Vicario, Michael A. Nitsche, Vahid Nejati

## Abstract

**Background:** The COVID-19 pandemic has spread worldwide, affecting millions of people and exposing them to home quarantine, isolation, and social distancing. While recent reports showed increased distress and depressive/anxiety state related to COVID-19 crisis, we investigated how home quarantine affected sleep parameters in healthy individuals.

**Methods:** 160 healthy individuals who were in home quarantine in April 2020 for at least one month participated in this study. Participants rated and compared their quantitative sleep parameters (time to go to bed, sleep duration, getting-up time) and sleep quality factors, pre-and during home quarantine due to the COVID-19 pandemic. Furthermore, participants’ chronotype was determined to see if sleep parameters are differentially affected in different chronotypes.

**Results:** The time to fall asleep and get-up in the morning were significantly delayed in all participants, indicating a significant circadian misalignment. Sleep quality was reported to be significantly poorer in all participants and chronotypes, and included more daily disturbances (more sleep disturbances, higher daily dysfunctions due to low quality of sleep) and less perceived sleep quality (lower subjective sleep quality, longer time taken to fall asleep at night, more use of sleep medication for improving sleep quality) during home quarantine.

**Conclusions:** Home quarantine due to COVID-19 pandemic has a detrimental impact on sleep quality. Online interventions including self-help sleep programs, stress management, relaxation practices, stimulus control, sleep hygiene, and mindfulness training are available interventions in the current situation.

## Introduction

In December 2019, the World Health Organization (WHO) announced the outbreak of a novel and severe respiratory virus called severe acute respiratory syndrome coronavirus 2 (SARS-CoV-2) in China (Wuhan) leading to the COVID-19 disease. Since then the virus and the resultant COVID-19 disease gradually spread throughout the world with more than 10 million confirmed cases and more than a half-million confirmed deaths in 216 countries and territories so far (WHO, retrieved: https://www.who.int/emergencies/diseases/novel-coronavirus-2019, as of June 30^th^ 2020). The disease was recognized as a substantial global public health emergency and a global pandemic was declared on March 11, 2020. As a result of the COVID-19 pandemic, strict restrictions, including quarantine, were declared by governments of seriously affected countries. This resulted in a significant and immediate change of lifestyle due to home-confinement situations (e.g. stay-at-home orders, quarantine, and isolation), mobile working, and restricted human-based communications (World Health Organization, 2020). These changes in lifestyle including quarantine and isolation have negative consequences for psychological health and wellbeing as shown in a recent review (Brooks et al., 2020).

In this line, recently published reports after the outbreak show negative impacts of the COVID-19 crisis on different aspects of psychological well-being and health (Pfefferbaum and North, 2020). Symptoms of anxiety and depression and self-reported stress have been common psychological reactions to the COVID-19 pandemic (Rajkumar, 2020) and are more severe in patients with mental health disorders and medical staff (Chen et al., 2020; Yao et al., 2020). These symptoms and negative states have been associated with sleep problems as well (Rajkumar, 2020) which are related to changes in sleep/wake cycle and circadian rhythms. Circadian rhythms are 24 h daily cycles that pervasively affect physiology and behavior in mammals, including humans. They can be entrained or phase-shifted not only by our internal clock but also external factors such as social stimuli, and daily schedules or social rhythms (Grandin et al., 2006). Any internal or external factor that deviates this cycle from its normal 24 h rhythm can lead to circadian misalignment and sleep difficulties. On the other hand, sleep quality is an important marker of mental health and well-being (Jike et al., 2018), and many immune parameters show systematic fluctuations over the 24□ h period in human blood (Scheiermann et al., 2013). The occurrence of sleep disturbances is frequently reported in a variety of psychiatric conditions (Breslau et al., 1996; Cole et al., 2006; Freeman et al., 2020), indicating an important role of sleep quality in humans well-being.

Home quarantine, mobile working, and restricted/limited social interactions due to the COVID-19 crisis can negatively affect sleep parameters and circadian alignment, as markers of psychological well-being and mental health. In the present study, we investigated quantitative sleep parameters, sleep quality factors, and circadian alignment in individuals during the time being in home quarantine for at least one month during the COVID-19 outbreak in Tehran, Iran in April 2020. The purpose of this study was to explore how quantitative parameters of sleep (e.g., time to go to bed, sleep duration, get-up time in the morning), sleep quality, and circadian rhythms are affected by home quarantine and lifestyle changes due to the COVID-19 crisis. We also investigated how individuals with different chronotypes (i.e., circadian preference) are differentially affected by home quarantine due to the COVID-19 crisis.

## Materials and methods

### Participants

One-hundred and sixty healthy volunteers (137 females, mean age = 25.79 ± 7.31) participated in this study. Participants were recruited online via university-based and non-university social media. The inclusion criteria were (1) being 18-60 years old, (2) being physically active in society, (3) and no reported psychiatric, psychological and neurological disorders, especially sleep-related disorders, at the time of recruitment. The study was conducted according to the latest version of the Declaration of Helsinki ethical standards and approved by the Ethical Committee of the Shahid Beheshti University.

### Measures

#### Pittsburgh Sleep Quality Index (PSQI)

The PSQI is a widely used 19-item self-report questionnaire for measuring sleep disturbances and healthy sleep (Buysse et al., 1989; Backhaus et al., 2002; Murawski et al., 2018). It includes seven clinically derived domains of sleep difficulties, including sleep quality, sleep latency, sleep duration, habitual sleep efficiency, sleep disturbances, use of sleeping medications, and daytime dysfunction. Each domain score is calculated based on response to specific items, most of them presented in a 0-3 Likert-type (with higher scores indicative of poorer sleep quality). These sleep domains are combined to a single PSQI Sleep Quality factor, with a higher score indicating worse sleep quality. In addition to the global PSCI factor, a validated three-factor model of the PSQI is proposed to assess disturbances in three separate factors of subjective sleep reports, including sleep efficiency, perceived sleep quality, daily disturbances (Cole et al., 2006). The Persian version of the PSQI has proven reliability, and validity with a Cronbach’s alpha of 89.6 (Ahmadi et al., 2010). The PSQI used in this study was structured to ask participants to answer each item for the time before home quarantine, and for the month during which they have been at home quarantine (see appendix 1 for an example).

#### Quantitative sleep parameters

Questions 1-4 of the PSQI ask about time to go to bed (clock), sleep onset latency time (in minutes), time to get up in the morning, and sleep duration. These questions were customized to obtain each parameter for the time before home quarantine, and for the month during which they have been at home quarantine. Responses were used as measures of quantitative sleep parameters.

#### Morningness-Eveningness Questionnaire

Chronotype was assessed by the Morningness-Eveningness-Questionnaire (Horne and Ostberg, 1976). The questionnaire consists of 19 questions that ask individuals to determine their “feeling best” rhythms, indicate preferred clock time blocks rather than the actual real time for sleep and engagement in other daily/weekly activities (e.g. physical exercise, tests, work), and assess morning alertness, morning appetite, and evening tiredness. Each question has a score and the sum scores range from 16 to 86, with scores below 42 indicating evening type or late chronotype, and scores higher than 58 indicating morning type or early chronotype. Definite evening (16-30), moderate evening (31-41), intermediate or neutral (42-58), moderate morning (59-69) and definite morning (70-86) are the five chronotype categories identified by the MEQ. The questionnaire shows high reliability and significantly correlates with circadian rhythm-related hormonal changes, including melatonin (Griefahn et al., 2001). In addition to the MEQ, we defined the subjective chronotype phenotype as the question “Are you naturally an evening person or a morning person or neither?” (Jones et al., 2019). The Persian version of the MEQ has high validity with a Cronbach’s alpha of 0.79 (Rahafar et al., 2013).

### Data collection

To prevent the spread of COVID-19 and comply with the ethical protocol of the ethical board, data collection was conducted online and via a web-based survey. The measures were designed for web-based use (Google Doc format) and sent to volunteers via social media. Participants were offered sleep hygiene packages including educational material for their participation after they completed the survey. Participants answered the questionnaires anonymously from April 20^th^ to April 28^th^ during a national call for home quarantine and were required to be in quarantine for at least 4 weeks before data collection.

### Statistical analysis

For quantitative sleep parameters (e.g. time to go to bed, sleep onset latency time, duration of sleep, time to wake up), paired sample t-tests (two-tailed, *p* < 0.05) were used to compare respective parameters before and during home quarantine. For statistical analysis of respective changes, the delay of “time to go to bed” was added to midnight, i.e. 24:00 (a person whose average time to going to bed moved from 24:00 to 2:00 AM scored at 26). For analysis of the sleep quality parameters, the global PSQI score obtained before the pandemic and during home quarantine was compared with a paired t-test. Furthermore, a within-subject 3 × 2 repeated-measures ANOVA was conducted on sleep quality-derived factors with sleep quality (sleep efficiency, perceived sleep quality, daily disturbances) and condition (pre vs during quarantine) as within-subject factors. In the case of significant results, post hoc comparisons were conducted with Bonferroni-corrected post hoc t-test. The potential confounding effects of gender and age on sleep parameters were investigated by including age and gender as potential covariates in an ANCOVA analysis. In addition to the global score and compound factors of sleep quality, each of the Likert-scaled PSQI items was compared before and during home quarantine using the Wilcoxon signed-rank test (*p* < 0.05). Finally, the association of chronotype and quantitative sleep parameters and chronotype and sleep quality were investigated by 3 × 2 and 3 × 3 mixed-model ANOVAs respectively with condition (2 values) or sleep quality factor (3 values) as the within-subject factors and chronotype (neutral, late, early) as the between-subject factor. In the case of significant results, post hoc comparisons were conducted with Bonferroni-corrected post hoc t-tests. For all ANOVAs, normality of the data and homogeneity of variance-covariance matrices were confirmed by the Kolmogorov-Smirnov and Levin tests, respectively. In case that the assumption of sphericity was violated in Mauchly’s test, the degrees of freedom were corrected using Greenhouse-Geisser estimates of sphericity.

## Results

### Quantitative sleep parameters

#### Time to go to bed, sleep onset latency, sleep duration, and getting up time

We had no missing data and all participants responses were used in the final analysis. 73.8 % of the participants (118 of 160) reported at least 1 h delay to go to bed during the pandemic situation after being in home quarantine for at least one month compared to pre-quarantine time. The average delay time to go to bed during quarantine was 1.48 h. To test for statistical significance, the time to go to bed pre-vs during quarantine was compared, and the results show that participants’ time to go to bed was significantly delayed during quarantine (*t* = 11.60, *p* < 0.001) (Fig. 1A). The average time taken for participants to fall asleep (in min) was compared as well and the results of the t-test showed that the time to fall asleep was prolonged during home quarantine as compared to the time before quarantine (*t* = 7.32, *p* < 0.001). The average sleep duration (in h) was significantly longer during quarantine, as compared to before quarantine as well (*t* = −3.65, *p* < 0.001) (Fig. 1B). Furthermore, 85.6 % of the participants (137 of 160) reported at least 1 h delay to get up in the morning on average in home quarantine. The average delay to get up in the morning during home quarantine was 2.28 h. Results showed that participants’ time to get up in the morning was significantly delayed during quarantine (*t* = 15.36, *p* < 0.001) (Fig. 1C).

**Fig 1:**
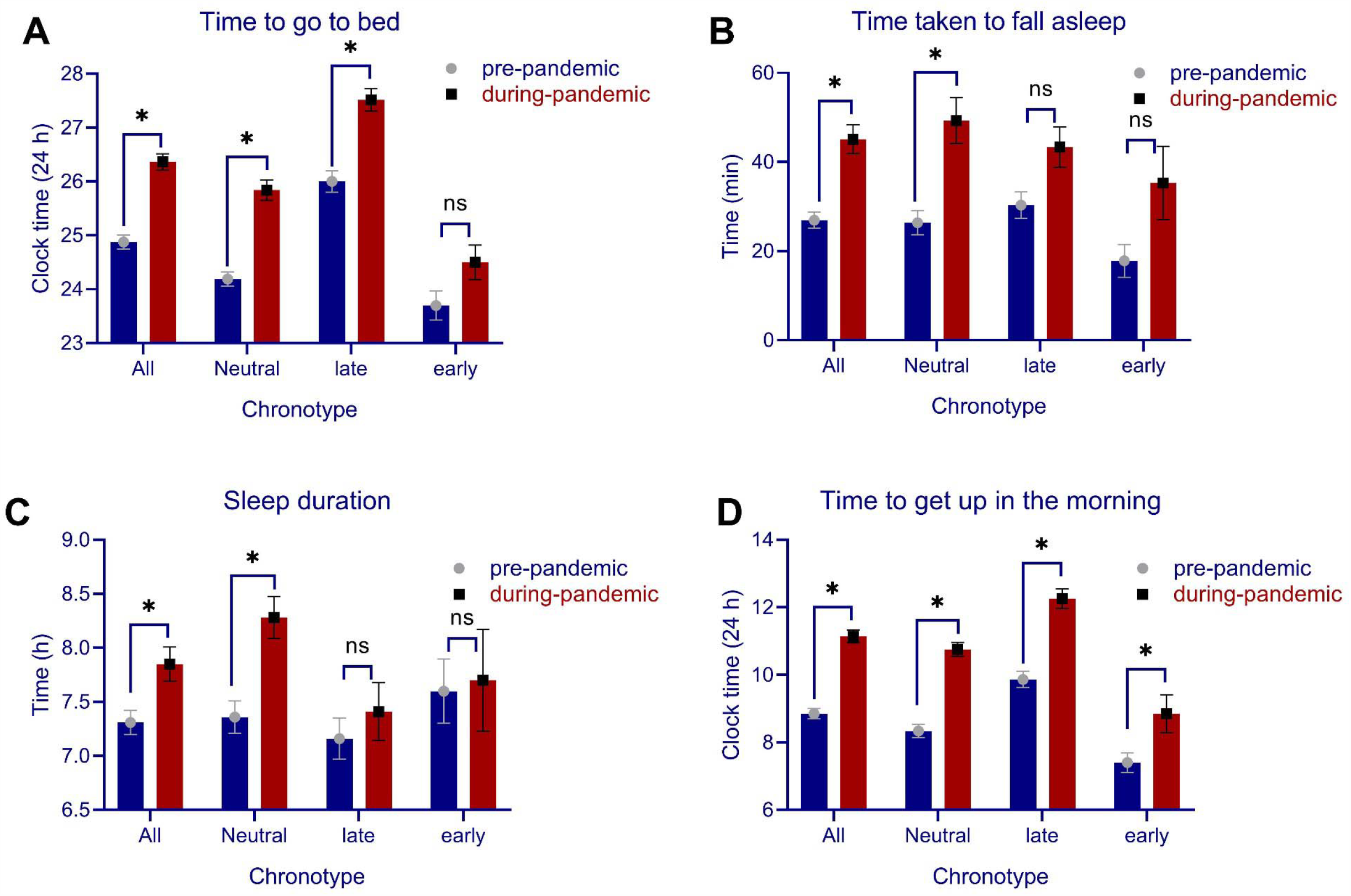
Quantitative sleep parameters of time to go to bed (**A**), time taken to fall asleep (in min) (**B**) sleep duration (**C**), and time to get up in the morning (**D**) in participants before the COVID-19 pandemic compared to one month after the COVID-19 pandemic during home quarantine. *Note*: In figure A, the time to go to bed after 24:00 is calculated as 24 + *n* where *n* stands for the time after 24:00. All error bars represent standard error of mean (s.e.m). h = hour. *n* = 160.

### Sleep quality

The global PSQI score, which serves as an index of general sleep quality, was analyzed before and after the pandemic crisis. The results of the paired t-test revealed a significant decrease of global sleep quality during quarantine (*t* = 6.95, *p* < 0.001). In addition to the global PSQI, the three suggested factors of sleep quality (sleep efficiency, perceived sleep quality, daily disturbances) (Cole et al., 2006) were analyzed before and after the pandemic crisis with a 3 × 2 ANOVA. The results of the ANOVA showed a significant interaction of condition × sleep quality factors (*F*_*2*_ = 29.07, *p* < 0.001, η_***p***_^*2*^ = 0.15) as well as significant main effects of condition (*F*_*2*_ = 48.41, *p* < 0.001, η_***p***_^*2*^ = 0.23) and quality factors (*F*_*2*_ = 141.19, *p* < 0.001, η_***p***_^*2*^ = 0.47). Bonferroni-corrected *post hoc* t-tests showed that the factors perceived sleep quality (*t* = 8.58, *p* < 0.001), and daily disturbances (*t* = 7.60, *p* < 0.001) were significantly lower and higher during home quarantine compared to pre-quarantine, whereas sleep efficiency was not significantly affected (*t* = 0.57, *p* = 0.565) (Fig. 2A,B). The potential confounding effects of gender and age were investigated in a separate ANCOVA, and results showed no interaction of condition with gender (*F*_*1*_ = 0.01, *p* = 0.935) and age (*F*_*1*_ = 0.79, *p* = 0.752). In addition, each individual domain of sleep quality was compared before and during quarantine. The results of the Wilcoxon signed-rank test conducted for each item showed a significantly lower subjective quality of sleep (*Z* = - 5.62, *p* < 0.001), longer time taken to fall asleep a night (*Z* = −4.92, *p* < 0.001), more sleep disturbances (*Z* = −3.52, *p* < 0.001), more use of sleep medication for improving sleep quality (*Z* = −3.69, *p* < 0.001) and finally significantly higher daily dysfunctions due to low quality of sleep (*Z* = −5.64, *p* < 0.001).

**Fig 2:**
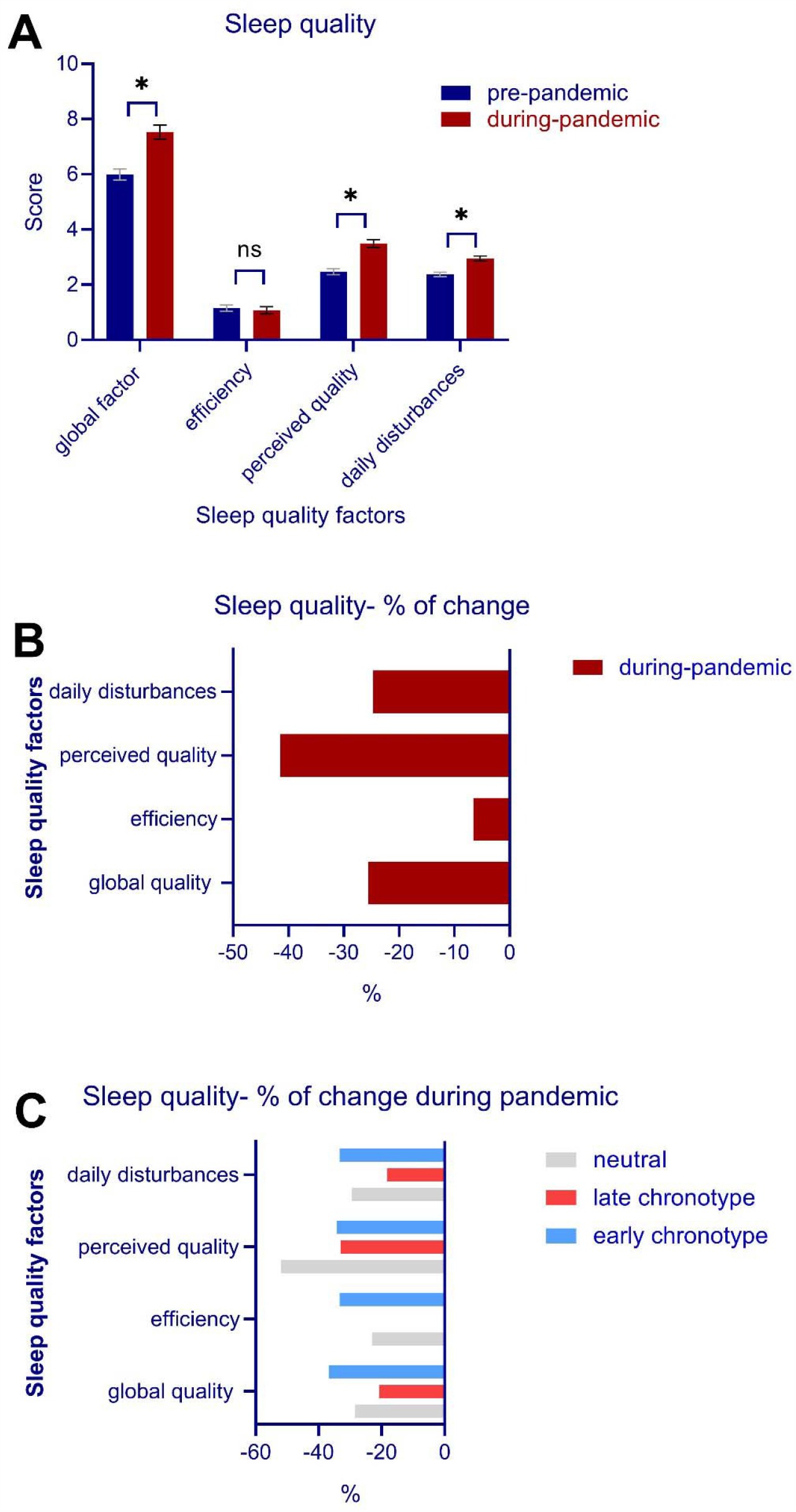
**A**, Sleep quality factors during the COVID-19 pandemic in home quarantine (**B**, percentage of sleep quality factors after the COVID-19 pandemic (**C**), percentage of sleep quality factors during the COVID-19 pandemic in participants based on chronotype.

### Chronotype, circadian alignment, and sleep quality

For quantitative sleep parameters, we analyzed the interaction of “condition” (pre vs during-pandemic time) and “chronotype” (neutral, late, early) for “time to go to bed” with a mixed-model ANOVA and found significant main effects of condition (*F*_*1*_ = 71.78, *p* < 0.001, η _***p***_^*2*^ = 0.31) and chronotype (*F*_*2*_ = 49.99, *p* < 0.001, η_***p***_^*2*^ = 0.38), but no interaction of condition × chronotype (*F*_*2*_ = 2.03, *p* = 0.134). Bonferroni-corrected *post-hoc* comparisons revealed that individuals with neutral (*t* = 6.63, *p* < 0.001), and late chronotype (*t* = 5.57, *p* < 0.001), but not early chronotype (*t* = 1.75, *p* = 0.246) had a significantly delayed in going to bed, indicating an alteration of their circadian-preferred time to go to sleep. With regard to getting-up in the morning, significant main effects of condition (*F*_*1*_ = 136.68, *p* < 0.001, η_***p***_^*2*^ = 0.46) and chronotype (*F*_*1*_ = 26.90, *p* < 0.001, η_***p***_^*2*^ = 0.25) were found, but condition did not significantly interact with chronotype (*F*_*2*_ = 2.20, *p* = 0.114). Bonferroni-corrected *pot hoc* t-tests revealed that all chronotypes, including neutral (*t* = 7.46, *p* < 0.001), late (*t* = 6.97, *p* < 0.001), and early chronotype (*t* = 2.44, *p* = 0.045) had a significant delay in getting up in the morning during home quarantine (Fig. 1A,D). In addition to time to go to bed and time to get up in the morning, which are most relevant for circadian alignment, we analyzed the potential impact of chronoytpe on the time taken to fall asleep and sleep duration as well. A mixed model ANOVA was conducted on time taken to fall asleep. The results showed no interaction of condition × chronotype (*F*_*2*_ = 1.75, *p* = 0.177) or main effect of chronotype (*F*_*2*_ = 1.22, *p* = 0.296), but the main effect of condition was significant (*F*_*2*_ = 5.98, *p* = 0.016). Similarly, for sleep duration, no interaction of condition × chronotype (*F*_*2*_ = 2.90, *p* = 0.058) or main effect of chronotype were found (*F*_*2*_ = 2.45, *p* = 0.089) but the main effect of condition was significant (*F*_*2*_ = 5.98, *p* = 0.016). This indicates that the longer time taken to fall asleep and longer sleep duration during quarantine was not to specific chronotype and occurred in all participants (Fig. 1B,C).

Finally, the relationship between chronotype (3 values) and sleep quality was investigated. The results of the 2 × 2 ANOVA showed no interaction of chronotype × condition on the global sleep quality factor (*F*_*1*_ = 0.16, *p* = 0.690). Furthermore, the association of chronotype, sleep quality factors (3 values) and quarantine condition (2 values) on sleep quality were investigated using a 3 × 3 × 2 ANOVA. The main effects of chronotype (*F*_*2*_ = 7.25, *p* < 0.001, η_***p***_^*2*^ = 0.08), sleep quality (*F*_*2*_ = 84.49, *p* < 0.001, η_***p***_^*2*^ = 0.35), condition (*F*_*1*_ = 33.05, *p* < 0.001, η_***p***_^*2*^ = 0.17), and the interaction of sleep quality × condition (*F*_*1*_ = 13.61, *p* < 0.001, η_***p***_^*2*^ = 0.08) were significant. Chronotype × sleep quality (*F*_*3*.*97*_ = 2.14, *p* = 0.079) and chronotype × sleep quality × condition (*F*_*3*.*76*_ = 2.08, *p* = 0.098) did not significantly interact, indicating that poor sleep quality did not depend on specific chronotype (Fig. 2C).

## Discussion

In the present study, we investigated quantitative sleep parameters, sleep quality factors, and circadian alignment in 160 individuals who were in at home quarantine and isolation for at least 1 month during the COVID-19 outbreak in April 2020. All quantitative sleep parameters were negatively affected. Specifically, the time to go to bed and time to get up in the morning were significantly delayed and the time needed to fall asleep, as well as sleep duration were significantly longer during quarantine. This circadian misalignment occurred in all chronotypes (neutral, late, early chronotype). Furthermore, participants reported a significantly poorer sleep quality in home quarantine during the COVID-19 crisis compared to the pre-quarantine time. These factors included more daily disturbances (e.g. more sleep disturbances, higher daily dysfunction due to low quality of sleep) and less perceived sleep quality (e.g. lower subjective sleep quality, longer time taken to fall asleep a night, more use of sleep medication for improving sleep quality) during the COVID-19 pandemic. Consequently, lower general sleep quality was observed in all chronotypes.

Circadian rhythms provide a temporal organization of physiological processes and behavior to promote effective adaptation to the environment. At the behavioral level, this is achieved via regular cycles of sleep and waking, and circadian misalignment is the consequence of irregular sleep/waking cycles (Robert and Moore, 1997). The change of quantitative sleep parameters during the COVID-19 crisis falls in the category of *delayed phase sleep disorder* (DPSD) which is the most prevalent circadian rhythm disorder, and is characterized by the inability to fall asleep and arise at conventional clock times. DPSD depends to a great extent on lifestyle (Robert and Moore, 1997; Baron and Reid, 2014). It has been linked to negative health behavior, including but not limited to higher smoking rates, and alcohol use (Saxvig et al., 2012), higher depressive symptoms (Abe et al., 2011), and higher prevalence of affective disorders (Abe et al., 2011; Lee et al., 2011) in previous studies. Although the delayed phase state reported in the participants during the COVID-19 quarantine might be transient if the situation changes back to routine, which was not explored in this study, a negative impact of circadian misalignment (acute or chronic) on health was established in previous studies (Baron and Reid, 2014). Emotional difficulties (e.g. emotional reactivity, emotional dysregulation) and increased levels of anxiety and stress are commonly associated with circadian misalignment. Furthermore, daily circadian disruption can impair cognitive performance and learning processes due to the regulatory role of the sleep-wakefulness cycle in modulating arousal, neurocognitive and affective functions (Wright et al., 2012; Chellappa et al., 2018). This has important implications for working and educational environments and could negatively affect performance of workers doing mobile working or students involved in online learning materials.

In addition to negative impacts on healthy individuals, circadian misalignment is typical for several clinical conditions, such as depression, bipolar disorders, and attention-deficit hyperactivity disorder (Grandin et al., 2006; Baron and Reid, 2014; Alloy et al., 2015). In bipolar spectrum disorders, for example, a social/circadian rhythm model has been proposed for the development and course of this disorder (Alloy et al., 2015). According to this model, life events that disrupt the social zeitgeber, which are defined as daily socially determined rhythms or schedules (e.g. bedtimes, mealtimes, start and end of school, or occupation), can precipitate bipolar symptoms, particularly involving somatic symptoms such as sleep changes, because they disturb circadian rhythms. The reported higher depressive and anxiety states in the general population as well as in patients with mental disorders during the COVID-19 crisis (Chen et al., 2020; Rajkumar, 2020; Yao et al., 2020) could be at least partially due to circadian misalignment. One of the major negative impacts of poor sleep involves emotional regulation. Recent studies show that sleep critically affects daytime mood, emotional reactivity and the capacity to regulate positive and negative emotions (Kamphuis et al., 2012; Gruber and Cassoff, 2014; Palmer and Alfano, 2017). At the behavioral level, poor sleep negatively affects emotions at various stages of the regulatory process, including the identification, selection, and successful implementation of various strategies to deal with emotions, and leads to decrements of motivation and perceived reward (Palmer and Alfano, 2017). These emotional dysregulatory effects are proposed to be caused by the interaction of the emotional centers of the brain with the homeostatic sleep system (Gruber and Cassoff, 2014), and increased neural excitability in the brain after poor sleep (Elmenhorst et al., 2017; Palmer and Alfano, 2017).

Besides quantitative parameters of sleep and circadian misalignment, sleep quality was reported to be significantly poorer during the COVID-19 crisis. This is reflected by a significantly *lower perceived sleep quality*, longer time needed to fall asleep, and higher use of medication to assist falling asleep. The other significantly impaired factor of sleep quality was *daily disturbance* which significantly increased in the post-pandemic time. This factor indicates higher daytime dysfunctions as a consequence of sleep disturbances, which can result in experiencing more frustration and negative emotions (Bower et al., 2010). Similar to circadian misalignment, poor sleep quality has been linked to increased negative emotions and reduced quality of life in healthy individuals (Bower et al., 2010; Choi et al., 2019), enhanced severity of symptoms in some psychiatric disorders (e.g., mood disorders, pain), and physical illness (Zhang et al., 2016; Becker et al., 2017; Lee et al., 2017; Castro-Marrero et al., 2018).

We investigated also the role of circadian preference or chronotype on the reported sleep parameters. Previous studies suggested that being late chronotype is a risk factor for poor general health, and experience of clinical conditions (e.g. depressive symptoms, obesity) compared to early chronotypes, presumably due to reduced lifestyle regularity (Baron and Reid, 2014). Our results showed however that all quantitative sleep parameters were similarly negatively affected in all chronotypes except for the time to go to bed, in which we found a trendwise, but no significant delay for early chronotypes, which should be carefully interpreted considering the low number of early chronotype participants in the sample. Similarly, sleep quality was worsened in all chronotypes in the pandemic situation, showing that sleep health was endangered in all individuals regardless of chronotype.

Overall, the results of the present study show poor sleep health (both sleep quality and quantity) in individuals being at quarantine and isolation during the COVID-19 pandemic compared to the time before the beginning quarantine. The majority of females in the sample, and low number of early chronotypes are limitations of this study, which need to be addressed in further studies with larger samples. Furthermore, our results are based on self-report data, and we did not obtain data about intervening psychological conditions which could at least partially account for the observed effects (e.g. anxiety, distress, depressive state). Nevertheless, the current data suggest a remarkable negative impact of the COVID-19 crisis, especially with respect to quarantine, on sleep health and indirectly on psychological health parameters. This calls for targeted interventions for improving sleep parameters, and prevention of associated psychological consequences. Available interventions for improving sleep health are stress management and relaxation practice, stimulus control, sleep hygiene, and exercise (Murawski et al., 2018). These interventions are feasible for home practicing during the COVID-19 crisis when treatment-seeking is not possible. Specifically, sleep hygiene education/programs could be useful for improving sleep health. Sleep hygiene programs include a set of behavior that improves sleep health, such as regular sleep schedules, and avoidance of long naps or the use of caffeine before sleep. These instructions can be trained via public media or offered as online interventions. Previous studies have shown good efficacy of internet-based sleep-promoting programs and mindfulness training for improving sleep quality (Suzuki et al., 2008; Querstret et al., 2017), which can be applied in the current pandemic situation. For those individuals that suffer from a joint clinical condition such as anxiety or mood disorders, interventions targeting cognitive and behavioral self-regulation are proposed for improving sleep quality (Murawski et al., 2018).

Commonly-used cognitive and behavioral interventions are stress management/relaxation, controlled breathing, and stimulus control (Murawski et al., 2018) which can again be instructed online in the current pandemic situation. Considering the unpredictability of the COVID-19 outbreak, which might extend for months, implementation of sleep health interventions is suggested, and should be taken into account by health authorities, policymakers and governments.

Appendix 1

## Instruction

The following questions relate to your usual sleep habits during the past month **before** the home quarantine due to COVID-19 outbreak and **during** the home quarantine due to COVID-19 outbreak:

## Question 2

a. When have you usually gotten up in the morning ***during*** the past month while being in home quarantine? ………….
b. When have you usually gotten up in the morning during the past month ***before*** the COVID-19 outbreak? ………….

## Data Availability

The datasets used and/or analyzed for the present study are available from the corresponding author upon reasonable request

---

**Table 1.** Mean and standard deviation of quantitative sleep parameters and sleep quality factors before and 1 month after the begin of the COVID-19 crisis in individuals being in quarantine

| Parameter | measure | Time | overall | chronotype |  |  |
| --- | --- | --- | --- | --- | --- | --- |
|  |  |  |  | neutral | late | early |
| Sleep quantitative | Time to go to bed<sup>1</sup> | pre-pandemic | 24.87±1.66 | 24.18±1.14 | 26.00±1.60 | 23.70±1.26 |
|  |  | during-pandemic | 26.36±1.94 | 25.83±1.63 | 27.51±1.69 | 24.50±1.50 |
|  | Sleep onset latency (min) | pre-pandemic | 26.94±23.19 | 26.39±23.48 | 30.32±23.94 | 17.80±17.15 |
|  |  | during-pandemic | 45.09±40.67 | 49.30±44.22 | 43.35±36.88 | 35.30±38.66 |
|  | Sleep duration (h) | pre-pandemic | 7.31±1.41 | 7.36±1.28 | 7.16±1.54 | 7.60±1.39 |
|  |  | during-pandemic | 7.87±1.99 | 8.28±1.66 | 7.41±2.17 | 7.70±2.20 |
|  | Time to get up | pre-pandemic | 8.85±1.98 | 8.37±1.68 | 9.86±1.96 | 7.40±1.39 |
|  |  | during-pandemic | 11.13±2.40 | 10.75±1.79 | 12.25±2.34 | 8.85±2.62 |
| Sleep quality | Global PSQI | pre-pandemic | 5.99±2.57 | 5.85±2.40 | 6.68±2.69 | 4.15±1.74 |
|  |  | during-pandemic | 7.52±3.26 | 7.51±3.21 | 8.07±3.38 | 5.68±2.28 |
|  | Sleep efficiency | pre-pandemic | 1.15±1.39 | 1.10±1.36 | 1.31±1.49 | 0.73±1.14 |
|  |  | during-pandemic | 1.07±1.60 | 0.85±1.37 | 1.31±1.83 | 1.10±1.59 |
|  | Perceived quality | pre-pandemic | 2.47±1.41 | 2.31±1.27 | 2.78±1.56 | 2.00±1.20 |
|  |  | during-pandemic | 3.49±1.76 | 3.51±1.74 | 3.71±1.85 | 2.68±1.29 |
|  | Daily disturbances | pre-pandemic | 2.37±1.00 | 2.43±0.98 | 2.57±0.94 | 1.42±0.76 |
|  |  | during-pandemic | 2.95±1.13 | 3.14±1.18 | 3.04±1.01 | 1.89±0.65 |

*Note:* 1= The time to fall asleep after 24:00 was calculated as 24+n where n is the time after 24:00 (AM). In PSQI, a lower score is indicative of healthier sleep. PSQI = Pittsburgh Sleep Quality Index.

